# Automatic Speech Recognition and Phonetics-Informed Sentence Design for Spastic Dysarthria Detection and Corticobulbar Lesion Localization

**DOI:** 10.64898/2026.06.02.26354698

**Authors:** Chayoot Marukatat, Keerati Kaewrak, Sedthapong Chunamchai, Chaipat Chunharas

## Abstract

Spastic dysarthria diagnosis through subjective neurologist auditory-perceptual assessment remains standard practice despite known inaccuracy. To address this gap, we developed an objective framework grounded in phonetic evidence that spastic dysarthria preferentially impairs initial consonant articulation, using automatic speech recognition (ASR) to quantify dysarthria and localize corticobulbar lesions. We created four reading sentences targeting groups of initial consonants: labial (facial), lingual-alveolar (tongue), and velopharyngeal (pharyngeal/soft-palate) sentence, along with a mixed-consonant sentence for comparative evaluation. Thirty-seven patients with neuroimaging-confirmed corticobulbar lesions and 37 controls read each sentence. ASR transcribed dysarthric speech into text, and we computed a ‘syllable-error score’ by counting incorrectly transcribed syllables. This yields a clinically meaningful feature that makes syllable-level phonetic errors explicit. Logistic regression models were trained for each sentence, and performance was summarized by the area under the receiver operating characteristic curve (AUC) across 10,000 resampled train-test splits. Consonant-specific sentences significantly outperformed the mixed sentence: the lingual-alveolar sentence performed best with (median AUC 0.88), followed by the labial (0.80), then the velopharyngeal sentence (0.72), while the mixed-consonant sentence was lowest (0.67). These results suggest that the interpretable ASR-derived syllable error feature, combined with a relevant machine learning classifier could inform clinical insight into consonant-specific vulnerability in spastic dysarthria, with lingual-alveolar consonants appearing particularly informative. Overall, this novel ASR-based framework, together with phonetics-informed feature design provides objective, accurate, and clinically meaningful digital quantification for spastic dysarthria detection and corticobulbar lesion localization.

**Author summary:** At the bedside, neurologists often detect dysarthria by listening to a patient’s speech, a practical but subjective approach that may delay diagnosis when dysarthria is mild. We developed a simple digital assessment approach that combines clinical phonetic knowledge with an accessible artificial intelligence tool, automatic speech recognition (ASR), to make the bedside judgement more objective while remaining clinically meaningful. Rather than relying on pre-existing open speech datasets, we newly collected speech recordings from patients with neuroimaging-confirmed corticobulbar lesions and matched healthy participants. Most patients had mild dysarthria, representing a setting where perceptual diagnosis is uncertain. We designed sentences that target different speech muscle groups and used ASR transcription errors as measurable features. A simple machine learning classifier was then trained on these features to evaluate diagnostic performance. The best sentence designs distinguished patients from controls with good performance and produced understandable results in relation to speech physiology. Our study illustrates a broader principle for digital neurology: artificial intelligence may be most useful when it is guided by clinical knowledge rather than replacing it and when it is available at the bedside. This approach could shift neurological assessment toward objective yet explainable way and could be extended beyond diagnosis to repeated monitoring during speech rehabilitation.

## Introduction

Spastic dysarthria is one major type of motor speech disorder characterized by imprecise consonant articulation, harsh and strained voice quality, and reduced speech intelligibility, with significant localizing value to corticobulbar tract lesions (1). This condition results from impaired muscular control of the speech mechanism due to central nervous system damage, leading to abnormalities in speed, strength, accuracy, range, tone, or duration of articulation (2). It often presents as an early symptom of stroke, with a reported prevalence of 25% to 41% (3, 4). Early detection of spastic dysarthria is crucial for accurate lesion localization, prompt neurological diagnosis, and effective therapeutic intervention.

The diagnosis of spastic dysarthria and its underlying lesion predominantly relies on subjective auditory-perceptual analysis. Studies have revealed that neurologists exhibit poor performance in detecting spastic dysarthria with detection rate of only 35-40% and an inter-rater reliability of less than 0.2 (5, 6). This inaccuracy is due to limited linguistic background, inadequate auditory practice in recognizing deviant dysarthric characteristics, and the absence of validated objective tool. This could lead to misdiagnosis of the underlying lesion (7), particularly when dysarthria presents as isolated symptom without other clinical clues.

Recent advances in understanding the phonetic aspect of spastic dysarthria have emerged, correlating with clinical knowledge of its pathophysiology. Sriwimon et al. investigated the speech characteristics in spastic dysarthria and found that initial consonants are the most affected phonemic group, compared to vowels or final consonants. The least accurate sounds of initial consonants are those produced by lingual(tongue) muscle action (or ‘lingual-alveolar placement’, e.g. phoneme /t/, /d/), followed by velo-pharyngeal muscle action (e.g. phoneme /k/, /ŋ/) (8, 9). This observation aligns with the known pathophysiology of corticobulbar lesions, which predominantly affects particular muscle groups compared to others (10, 11). However, neurologists typically diagnose dysarthria through spontaneous speech during clinical encounters rather than using phonetically designed test sentences.

The present study aimed to develop a practical tool for the diagnosis of spastic dysarthria as well as detecting the causative brain lesion based on prior clinical and phonetic knowledge. We first developed four different consonant-based reading passages for both dysarthric and healthy participants. We used Automatic Speech Recognition (ASR) software to detect and quantify sentence errors (12), replacing subjective perceptual analysis by neurologists. The diagnostic power in detecting lesion in spastic dysarthria in terms of receiver operating characteristic (ROC) curve with area under the curve (AUC), sensitivity, and specificity involving each sentence-based predictive model was also assessed. The result indicated that the highest median AUC from the clinically guided model was 0.88 which is considered good and reliable. To the best of our knowledge, this is the pioneer proof-of-concept study aimed at innovating the practice of dysarthria lesion identification.

## Materials and Methods

### Study design and participants

This diagnostic cross-sectional case-control study was conducted at King Chulalongkorn Memorial Hospital, a comprehensive tertiary medical facility in Thailand. The study protocol was approved by the Institutional Review Board of Faculty of Medicine, Chulalongkorn University (reference IRB No. 0494/65 and COA No. 1202/2022), and all procedures were conducted in accordance with institutional ethical standards.

From July 2024 to December 2024, we recruited patients from both hospital inpatient and outpatient settings who had confirmed unilateral or bilateral corticobulbar lesion by brain CT/ MRI scan reviewed and reported by a radiologist with over 10 years of experience. Every patient had a definite neurological diagnosis, which was given by board-certified neurologist, related to the corticobulbar lesion. The dysarthric speech was inarguably perceived by both patient and caregiver and could be attributed specifically to spastic dysarthria due to the lesion location. Other inclusion criteria were participants with age ≥ 18 years and Thai ethnicity with native accent. Patients with impaired cognition including aphasia and other all-cause voice disturbances (e.g. hoarseness) were excluded.

Control participants were healthy participants with neither history of speech abnormality nor neurological disease from healthy aging clinic. Of note, control participants were selected in sex- and age-matched manner, as these factors can potentially influence the speech characteristics.

Total number of 74 participants including 37 patients and 37 control participants, were recruited (see Fig. 1). The research protocol was revealed and informed consent was obtained by the participant prior to the enrolment. Participants were then appointed for a recording session which was organized in sound-treated audiometric booth. Each participant was instructed to repeat after four phonetically-different Thai sentences. The speech recording was done by iPhoneXR’s microphone via Voice Memo™ application. The file was automatically converted to digital format and confidentially stored as audio file type ‘.m4a’ by Apple software. Final data of each participant consisted of four audio files. Additional clinical information including demographic data, neurological diagnosis, symptom onset, imaging findings, and dysarthria severity, subjectively evaluated by a speech-language pathologist using ‘*Intelligibility Score in Thai Dysarthria*’ (see S1 Fig), was recorded at the time of enrolment.

**Fig. 1.**
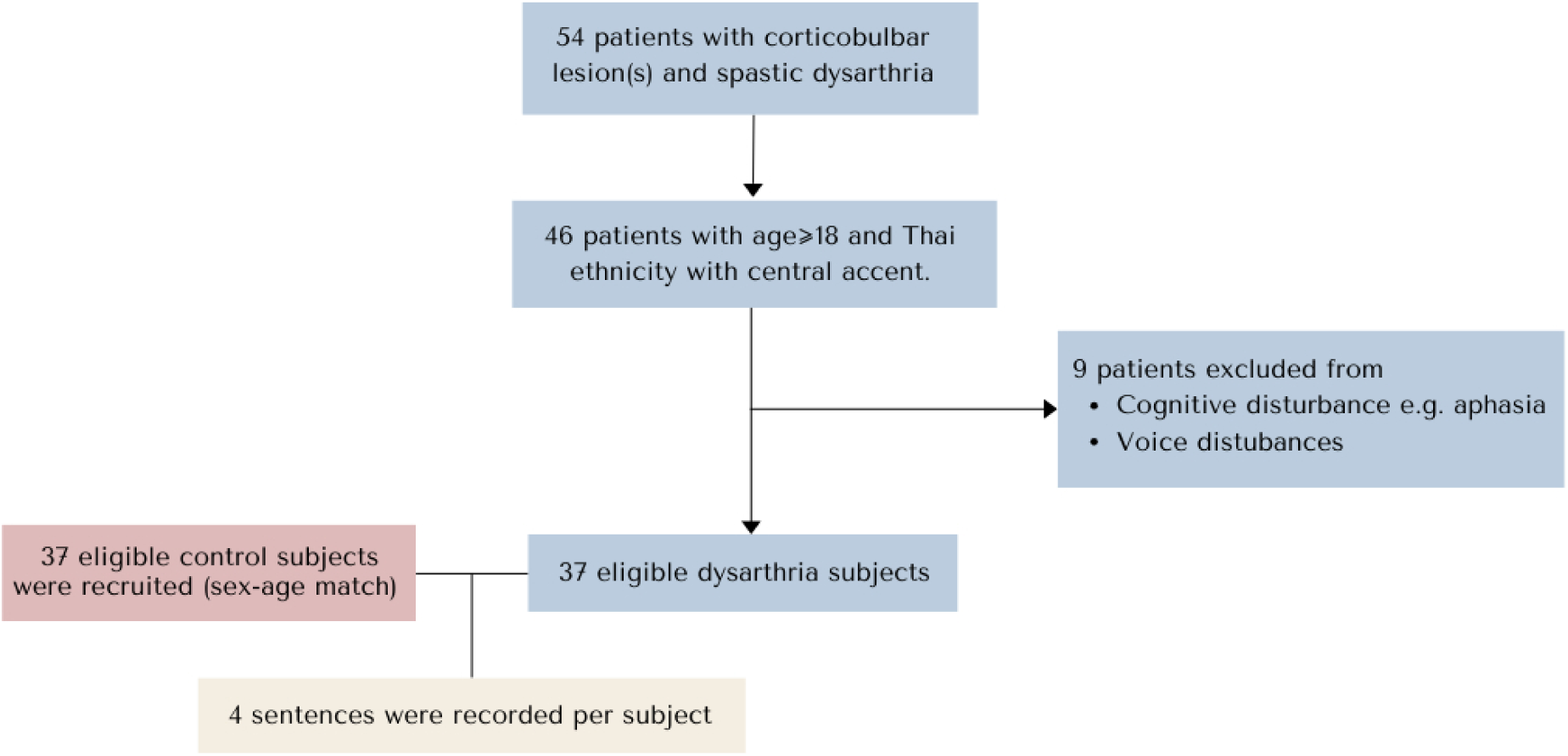
Flow diagram of participant recruitment. Figure shows a total of 54 dysarthria patients were screened, with 37 meeting eligibility criteria. An additional 37 age- and sex-matched controls were recruited. Each participant completed a single recording session with four sentences.

### Phonetics background-based sentence development (Feature design)

Findings from various studies in phonetics and phonology of spastic dysarthria indicated that initial consonant significantly deviated as compared to other syllable constituents (vowel, final consonant), in addition, some specific phonemes, the smallest sound unit in language, were affected more than others (7, 8). This implies that specific bulbar muscles are more likely to be impaired than others from corticobulbar lesion. As a result, articulating with these muscles would show significant imprecise syllable.

In collaboration with a speech-language pathologist, we developed four sentences featuring different groups of initial consonant phonemes, each articulated by a specific group of muscles. Each sentence was grammatically correct and meaningful in the Thai language and shared similar distribution of vowel sounds (front, central, back, as well as close, mid, open vowels) and with a comparable intonation level (low, mid, high, fall, and rising intonation).

1. The first sentence /pū: - má: - wîŋ - pāj - mā: - bōn - bāj – máj/ (Thai script: ‘ปูม้าว่ิงไปมาบนใบไม้’; meaning: ‘The crab runs back and forth on the leaves’), referred to as the Labial Placement (LBP) sentence, includes phonemes /p/, /b/, /m/, and /w/, which are primarily articulated using the lips. These labial phonemes engage facial muscles innervated by the facial nerve.
2. The second sentence /tɕʰāːw – râj – tàt - tôn - sǒn - tʰām - tʰɔ^ːn - sūŋ/ (Thai script: ‘ชาวไร่ตัดต้นสนทำท่อนซุง’; meaning: ‘The farmer cuts down pine trees to make logs’), referred to as the Lingual-Alveolar/Palatal Placement (LAP) sentence, includes phonemes such as /tɕʰ/ (ช), /r/ (ไร่), /t/ (ตัด), /tʰ/ (ทำ), and /s/ (สน), which involve different tongue placements. The /s/, /r/, /tʰ/ and /t/ sounds are articulated with the tongue against the alveolar ridge and teeth, while the /tɕʰ/ is a palatal sound. The /r/ is a drill sound. These movements are controlled by the hypoglossal nerve, which regulates tongue muscle actions.
3. The third sentence /ʔīː - kāː - kʰɔːj - kʰâːp - ŋūː - kʰâːp - kàj/ (Thai script: ‘อีกาคอยคาบงูคาบไก่’; meaning: ‘The crow is determined to carry a snake and a chicken’), named as the Velo-Pharyngeal Placement (VP) sentence, includes velar and pharyngeal sounds like /k/, /kʰ/, and /ŋ/, which involve pharyngeal and posterior tongue muscle contraction against the velum, innervated by the glossopharyngeal and vagus nerve. The glottal stop /ʔ/, produced by the momentary closure of the vocal folds, is controlled by the vagus nerve.
4. Final sentence /ph𝑖:aŋ – khæ: - fǒn – tòk – lōŋ - th𝑖𝑖 – nâ: - tà:ŋ – nāj – bā:ŋ - k^h^rā:’/ (Thai script: ‘เพียงแค่ฝนตกลงที่หน้าต่างในบางครา’; meaning: ‘Just when the rain sometimes falls on the window’, *trimmed lyrics of famous Thai song ‘Before rain’*), which expresses phonemes /ph/, /b/, /f/, /t/, /th/, /l/, /n/, /k^h^r/, /kh/, is referred to as Mixed Placement (MP) sentence. The sentence represents combined bulbar muscle action without selective features.

The development of these four sentences aimed to achieve two key objectives. First, to examine whether classification models using consonant-specific sentences (LBP, LAP, or VP) offer superior diagnostic performance compared to a non-selective mixed-consonant sentence (MP). Second, to determine which sentence provides the most accurate classification, thereby identifying the consonant group most susceptible to dysarthric impairment in spastic dysarthria.

### Feature extraction as syllable error score

After completing clinically guided feature design through the sentence development and recording, we quantified the dysarthric syllables by using the Artificial Intelligence tool instead of the expert auditory-perceptual analysis. The tool is known as Automatic Speech Recognition Software (ASR), in which the algorithm was based on Speech and Natural Language Processing deep learning model.

For each participant, four voice recordings were played through a speaker into the microphone of an iPhone XR running Apple Siri™ ASR. The ASR transcribed the speech into text, producing four written sentences per participant. Then we segmented each sentence into separate syllables. The transcribed data was organized and stored in a spreadsheet format for subsequent analysis (see S1 Table).

Syllables that deviated significantly from its intended pronunciation were transcribed incorrectly. Misspellings that altered the intended pronunciation, including incorrect initial, cluster, or terminal consonants, were counted as errors. Transcription errors in each sentence were quantified by simply counting syllable misspellings using the Python Pandas library, resulting in a ‘syllable error score’ per sentence. Individual participant was assigned with four error scores which served as input features for predictive model construction.

### Statistical analysis

Continuous variables following a normal distribution were presented as means with standard deviations, while variables with non-parametric distribution were described as medians and interquartile ranges. Categorical variables were reported as percentages. The normality of syllable error scores was assessed by Shapiro-Wilk Test which revealed that most scores followed a non-parametric distribution (see S2 Table). Subsequently, differences in error scores between case and control groups were analyzed with the Mann-Whitney *U* test (Wilcoxon rank-sum test), considering *p*-values < 0.05 (one-sided) as indicative of statistical significance. Categorical variables were evaluated using Chi-square test or Fischer’s exact test. Statistical analysis was performed with IBM SPSS statistics (version 29.0.1.0; IBM corporation, Armonk, NY, USA), and Python (version 3.10.12) using SciPy library (https://scipy.org).

### Dysarthria detection

#### 1. Data organization

We annotated the obtained error scores into ‘case’ if derived from a dysarthric patient with evidence of lesion, or ‘control’ if from a control participant. We then partitioned our dataset into training, validation, and test datasets according to standard protocol to improve model generalization. Initially, the data was randomly divided with a 70:30 train-test ratio, ensuring a stratified 1:1 distribution of case-control samples. To minimize sampling bias, this train-test division was resampled with different random states for 10,000 iterations. Within each iteration, the training set was shuffled and further split into training and validation datasets through stratified 5-fold cross-validation, using Python’s Scikit-learn library (https://scikit-learn.org/1.5/modules/generated/sklearn.linear_model.LogisticRegression.html), to validate model performance during training. The test set was held out to be unknown testing data for final evaluation.

#### 2. Model training

We trained four logistic regression models to detect dysarthria based on error scores obtained from each sentence (specifically LBP, LAP, VP, and MP sentence). In addition, multiple logistic regression model was also trained using four features from all sentences combined as predictor variables, referred to as ‘combined’ sentence model, to evaluate any improvement in classification performance. Training was conducted with maximum iterations of 100 using grid search for hyperparameter tuning to identify the optimal regularization parameter (C) which includes L2 regularization to prevent overfitting (13). The grid includes values from 10^-5^ to 10^5^, covering a range from strong regularization to weak regularization. A total of 10,000 iterations were conducted yielding five models per iteration. The model training was implemented using Python’s Scikit-learn library on Google Colab Jupyter Notebook environment without GPU support.

#### 3. Model evaluation and statistical analysis

To evaluate the performance of the model for each sentence feature, we primarily focused on AUC value which is the area under the receiver operating characteristic (ROC) curve. We assessed distributional assumptions for AUC per model using Shapiro-Wilk test. Because every AUC distribution deviated from normality, we summarized performance with median and interquartile range (IQR) and visualized the full nonparametric distributions using violin plots (see Fig. 3).

Pairwise differences in AUC between individual sentence models (LBP, LAP, VP, MP; excluding the combined-sentences model) were tested using one-sided Man-Whitney U tests. To control the family-wise error rate across the six pairwise comparisons, we applied a Bonferroni correction (α = 0.05/6), and report significance after correction in Fig. 3.

For each sentence, we selected the representative model whose AUC was closest to the median AUC across the 10,000 iterations. On this representative model, we reported sensitivity, specificity, precision, and F1-score at the optimal cutoff value defined by the Youden index.

#### 4. Permutation test

To confirm that model performance was significantly better than random chance, we conducted a permutation test. Participant labels (‘case’ and ‘control’) were randomly reassigned while preserving group sizes, and models were retrained using the same train-test split and training procedures as described above. This process was repeated for 10,000 times to generate the null distribution of AUC values for each model. The *p*-value was defined as the proportion of permuted AUCs equal to or greater than the observed median AUC (see S2 Fig).

## Results

Our first achievement was developing four novel Thai sentences, manually designed to capture the deviant characteristics of spastic dysarthria. The first three sentences were constructed to target specific groups of initial consonants based on the articulation muscles involved (consonant-specific sentences). In contrast, the Mixed Placement (MP) sentence incorporated multiple consonant groups (mixed-consonant sentence). The sentences were as follows (presented in phonemic transcription):

1. **pū: - má: - wîŋ -** pāj - mā: - bōn - bāj – máj (Labial Placement/LBP; facial muscles)
2. tɕʰāːw – râj – tàt - tôn - sǒn - tʰām - tʰɔ^ːn - sūŋ (Lingual-Alveolar/Palatal Placement/LAP; tongue muscles)
3. ʔīː - kāː - kʰɔːj - kʰâːp - ŋūː - kʰâːp – kàj (Velo-Pharyngeal Placement/VP, pharyngeal muscles)
4. ph𝒊:aŋ – khæ: - fǒ**n – tòk – lōŋ - th**𝒊𝒊 **– nâ: - tà:ŋ – nāj – bā:ŋ - k^h^rā:** (Mixed Placement/MP, combined muscles)

### Participant demographics

To ensure that the speech abnormalities were solely attributable to corticobulbar lesions, we matched participants in both groups by sex and age during recruitment, as these variables are the most significant contributors to the difference in speech characteristics. The mean age of the dysarthria and control group was 65.4 and 62.7 years old (p-value >0.05) respectively. Proportion of male sex was 78.4% and 73.0% (p-value >0.05) respectively. Thus, there were no statistically significant difference in sex and age between two groups.

The representativeness of the samples for spastic dysarthria was further demonstrated by etiological and lesion-specific diagnostic details, as illustrated in Table 1. All patients had confirmed corticobulbar lesion. Imaging studies revealed that 54.1% of lesions were located on the left corticobulbar tract, 35.1% on the right side and 10.8% bilaterally. The etiological diagnosis was mainly stroke (86.5%), followed by brain mass (10.8%) and demyelinating disease (2.7%). These anatomical and physiological findings provide robust evidence supporting the representativeness of spastic dysarthria in the patient samples.

**Table 1.**
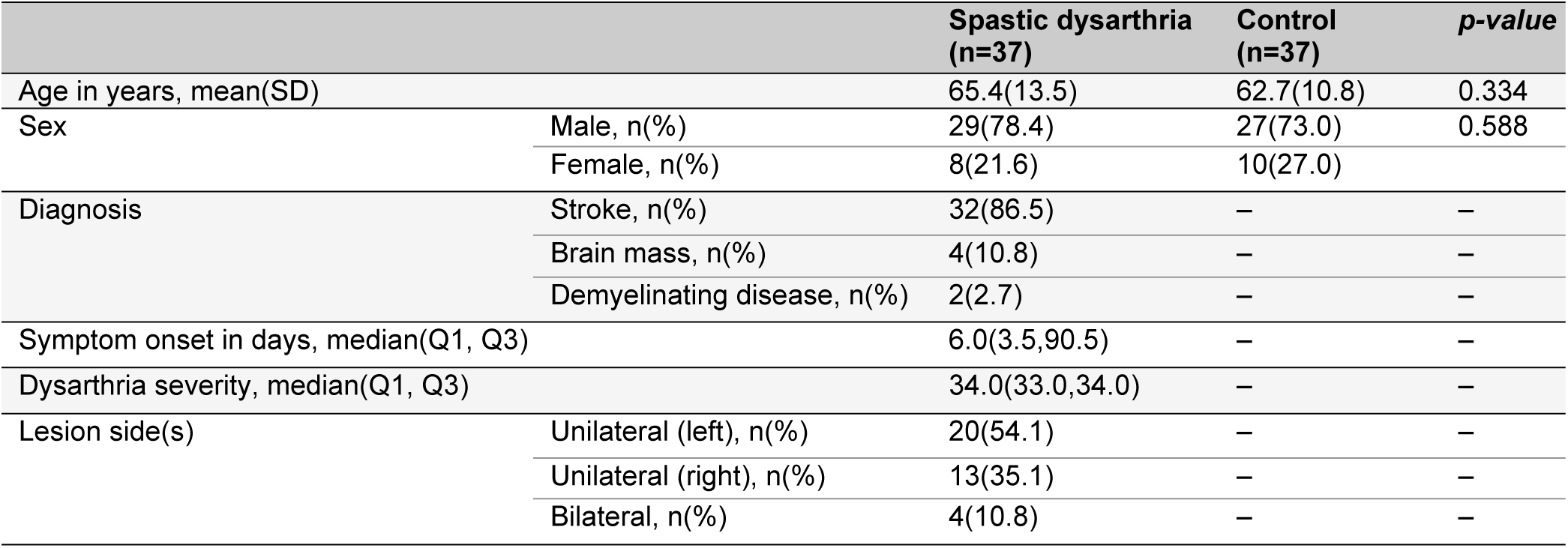
Demographic data between dysarthria and control group. Table summarizes age and sex for both groups, and primary diagnosis, dysarthria onset, dysarthria severity, and lesion side for the dysarthria group only. Dysarthria severity was evaluated using ‘*the Intelligibility Score in Thai Dysarthria*’, a subjective measurement in which higher scores indicate lower severity (maximum score = 36).

Regarding the perceptual classification of dysarthria severity, the median score of dysarthria severity was 34 (out of 36) with an interquartile range of 33 to 34. Higher scores indicate the lower severity, suggesting that most patients exhibited mild dysarthria with minimal impact on speech intelligibility. This highlights that the patient samples predominantly represented cases of mild dysarthria.

### Difference in syllable error scores for each sentence

‘Syllable error scores’ for each sentence, derived by transcribing speech from 74 participants (37 dysarthria and 37 controls) using Automatic Speech Recognition (ASR) followed by a count of incorrectly transcribed syllables, were primarily analyzed to determine whether there was a statistical difference between the dysarthria and control groups. The distributions of error scores were non-parametric and exhibited similar shapes across all sentences (see S2 Table).

Figure 2 presents the median error scores for each sentence. The median error score for the first Labial Placement (LBP) sentence was 2 (IQR 1-5) in the dysarthria group and 0 (IQR 0-1) in the control group (*U* = 283.0, *p* = <.001), indicating a statistically significant difference in error score. For the second Lingual-Alveolar/Palatal Placement (LAP) sentence, the median score was 3 (IQR 2-6) in the dysarthria group and 0 (IQR 0-1) in the control group (*U* = 175.0, *p* = <.001). The Velo-Pharyngeal Placement (VP) sentence had a median score of 4 (IQR 3-5) in the dysarthria group and 3 (IQR 2-3) in the control group (*U* = 390.5, *p* = 0.001). Lastly, the Mixed Placement (MP) sentence had a median error score of 1 (IQR 1-3) and 1 (IQR 1-1) in the dysarthria and control group respectively (*U* = 463.0, *p* = 0.003).

**Fig. 2.**
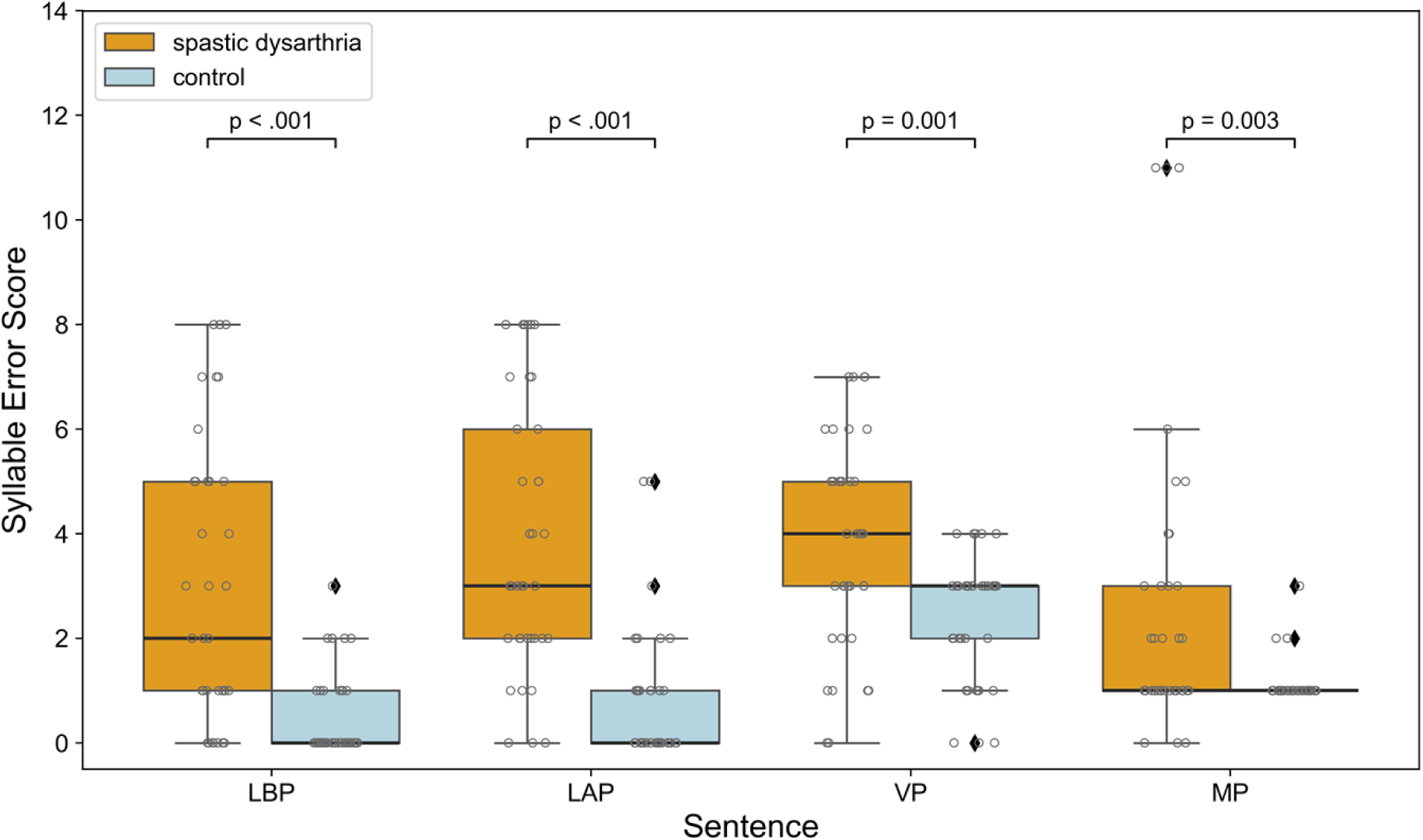
Scatter box plots of syllable error scores for each sentence in the spastic dysarthria and control groups. The sentences include Labial Placement (LBP), Lingual-Alveolar/Palatal Placement (LAP), Velo-Pharyngeal Placement (VP), and Mixed Placement (MP). The spastic dysarthria group consistently exhibited higher median error scores compared to the control group. All sentences showed statistically significant differences between the groups, with p-values indicated above each plot. Individual data points are overlaid as scatter plots to visualize participant-level variability.

These findings demonstrate consistent differences in syllable error scores across all sentences, indicating that Automatic Speech Recognition (ASR) can effectively quantify dysarthria and distinguish it from normal speech, while underscoring the potential of these error features for an accurate predictive model (For detailed Mann-Whitney U test statistics, see S3 Table).

### Predictive model

To predict spastic dysarthria lesion based on the syllable error scores of each sentence, four logistic regression models were trained using 5-fold cross-validation and evaluated against the unseen test dataset. Additionally, we developed a ‘combined’ sentence model that incorporated features from all sentences as predictor variables. Model performance was summarized as the median of area under the ROC curve (AUC) obtained from 10,000 iterations of randomized train-test data sampling. Figure 3 presents the non-parametric distribution of AUC scores for each model across all iterations. Consonant-specific models from LBP and LAP sentence, along with the ‘combined’ sentence model, demonstrated good performance, with median AUCs at 0.80 (IQR 0.75-0.84), 0.88 (IQR 0.83-0.92), and 0.89 (IQR 0.85-0.93), respectively. Among consonant-specific models, VP sentence showed lower performance with a median AUC at 0.72 (IQR 0.66-0.78). In contrast, the mixed-consonant model from MP sentence exhibited poor discriminative performance with median AUC at 0.67 (IQR 0.61-0.72) (see Table 2). These findings confirm that logistic regression classifier efficiently distinguishes between normal speech and dysarthria, when trained with selected phonetic features.

**Fig. 3.**
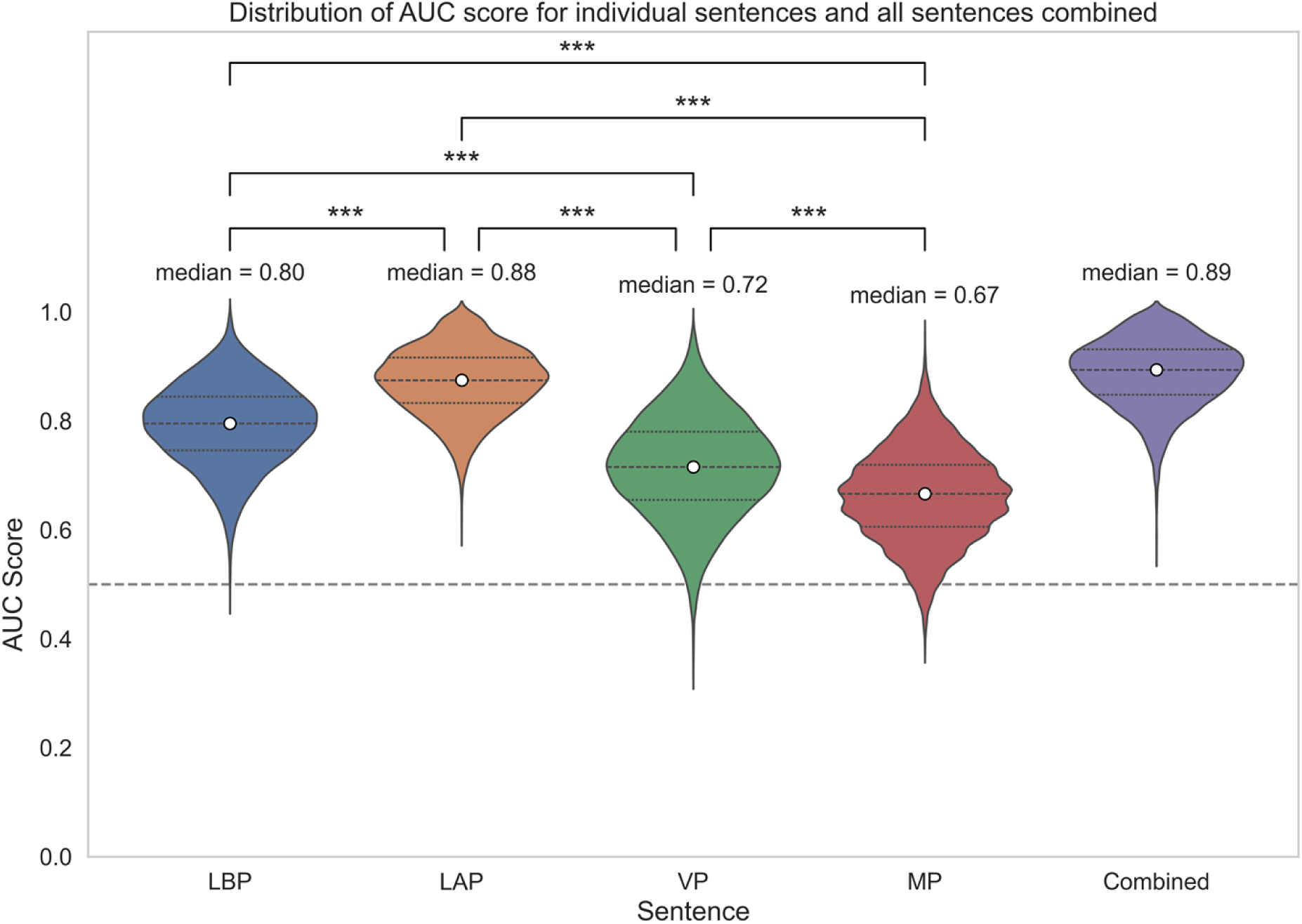
Distribution of Area Under the ROC curves (AUC). Violin plot showing the distribution of test AUC scores across 10,000 resampled splits with corresponding median AUC for models based on individual sentences and the ‘combined’ sentence. Statistical significance annotations indicate the significant difference in AUC between all pairwise models based on individual sentences. LBP = Labial Placement, LAP = Lingual-Alveolar/Palatal Placement, VP = Velo-Pharyngeal Placement, MP = Mixed Placement

**Table 2.**
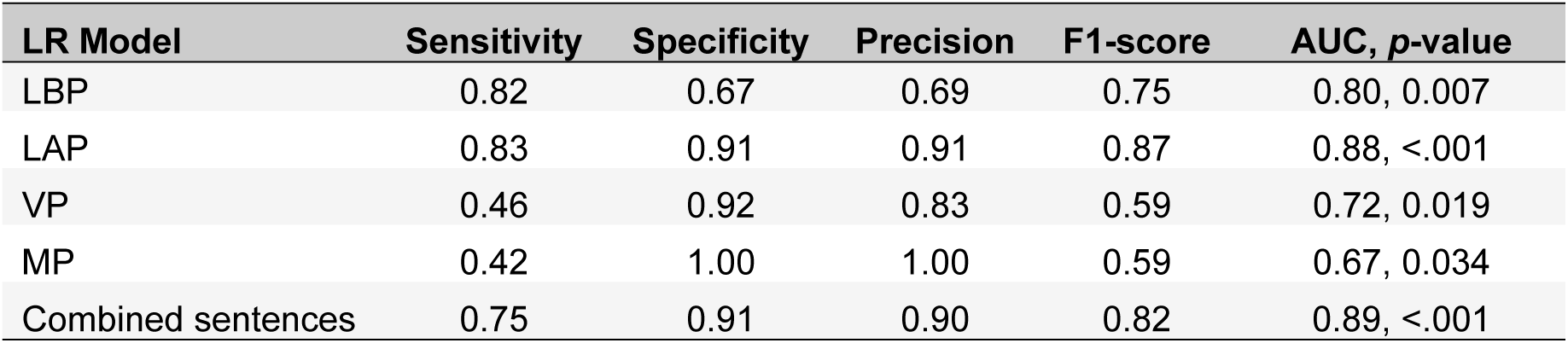
Performance of different models. Table shows performances of five representative logistic regression (LR) models for spastic dysarthria lesion detection based on individual sentences and ‘combined’ sentence, at the optimal threshold (Youden index).

Pairwise comparisons between models trained on individual sentences further highlighted the advantage of consonant-specific features. All consonant-specific models (LAP, LBP, and VP) showed statistically significant difference from mixed-consonant model (MP model), as well as showing significant differences among themselves (see Figure 3). The LAP model achieved a significantly higher AUC than the LBP model (*U* = 21.05×10^6^, *p* <.001), VP model (*U* = 92.18×10^6^, *p* <.001), and MP model (*U* = 97.75 ×10^6^, *p* <.001). the LBP model also outperformed the VP (*U* = 74.50×10^6^, *p* <.001) and MP models (*U* = 88.00×10^6^, *p* <.001), while the VP model remained significantly superior to the MP model (*U* = 67.10×10^6^, *p* <.001). These differences reflect the varying diagnostic value of different consonant groups.

Evaluation metrics were derived from the representative model whose AUC was closest to the median AUC across iterations, with *p*-values computed via permutation test. The LAP sentence-based model with the highest performance (AUC= 0.88, *p* <.001), achieved a sensitivity and specificity of 0.83 and 0.91, respectively, along with the precision of 0.91 and an F1-score, which balances sensitivity and precision, of 0.87, indicating strong predictive reliability. The LBP model (AUC = 0.80, *p* = 0.007) showed a sensitivity of 0.82 and specificity of 0.67, while the precision was 0.69 and an F1-score of 0.75. The VP model (AUC = 0.72, *p* = 0.019) relatively underperformed in sensitivity at 0.46 despite high trade-off specificity of 0.92, with an F1-score of 0.59. As expected, the MP model, with the lowest AUC (AUC = 0.67, *p* = 0.034), had poor sensitivity of 0.42 and specificity of 1.00. The ‘combined’ sentence model performed comparably to the LAP model (AUC = 0.89, *p* <.001) with sensitivity of 0.75, specificity of 0.91, and F1-score of 0.82. These results again demonstrate that consonant-specific sentences outperform mixed-consonant sentences in diagnostic accuracy (see Table 2).

## Discussion

In this study, we introduced a practical clinical quantification framework to augment subjective diagnosis and evaluated its effectiveness. Our results demonstrate two elements as critical to diagnostic performance: phonetic- and physiologic- informed sentence design and objective quantification of spastic dysarthria. Implemented via ASR-derived features and an interpretable logistic-regression classifier, this prior-knowledge feature design materially improves detection of spastic dysarthria. AUC distributions demonstrated consistently higher performance for the consonant-specific sentences (LAP, LBP, and VP), with the best models reaching AUCs up to 0.88, whereas the mixed-placement sentence (MP), designed without a targeted phonetic rationale, showed poor performance (median AUC = 0.67). Pairwise nonparametric comparisons corroborated these differences. Given that reported neurologist classification accuracy is typically below 50% (5, 6, 14), these findings underscore that linguistically and clinically informed feature design, more than added algorithmic complexity, is a key driver of diagnostic performance. Overall, these results support the practical utility of this framework for dysarthria detection and lesion localization.

A key clinical insight from this study is the central role of consonant articulation in detecting spastic dysarthria. Consistent with Sriwimon et al. (8), consonants articulated with lingual muscles with alveolar placement were most affected, as shown by the LAP sentence, which had the largest between-group difference in median syllable-error scores together with the highest diagnostic performance (see Fig. 2 & Fig. 3). Labial consonants articulated by facial muscles (LBP) also exhibited a significant between-group syllable-error difference with correspondingly strong diagnostic performance, consistent with clinical evidence of facial muscle involvement in stroke (2). These results imply that lingual-alveolar consonants could be the best candidate marker for dysarthria lesion identification–not necessarily because these muscles are preferentially affected by corticobulbar lesions, but because the sounds they produce are more susceptible to dysarthric impairment. Articulating consonants from a single muscle group within a sentence enhances the ability to distinguish dysarthric from normal speech, whereas a mixed-consonant sentence reduces classification performance. More broadly, these findings illustrate how clinically informed sentence/feature design, paired with ASR-transcripts that make syllable-level error visible, can support digital dysarthria quantification to deliver clinically interpretable insights.

Building on these findings, our approach has the potential to refine traditional bedside assessments for spastic dysarthria by replacing random speech evaluation with consonant-focused sentence testing. Threshold-based metrics (see Table 2) suggest distinct diagnostic roles for each sentence. The ‘Lingual-Alveolar Placement’ (LAP) sentence delivered a balanced profile between sensitivity (0.83), specificity (0.91), and F1-score (0.87), supporting both screening and diagnosis. Labial placement (LBP) achieved high sensitivity (0.82) but moderate specificity (0.67), indicating initial screening role. In contrast, the VP sentence showed lower sensitivity (0.46) but high specificity (0.92), suggesting a rule-in role. In practice, although these metrics were derived from representative models at Youden optimized cut-off, threshold can be tuned to screening versus confirmation priorities.

The cohort in this study focused on participants with predominantly mild dysarthria, as rated by a perceptual intelligibility assessment by a speech-language pathologist— cases that pose a diagnostic challenge in clinical practice, often leading to underdiagnosis of corticobulbar lesions. A representative model achieved an AUC of 0.89 for detecting subtle dysarthria, suggesting that our proposed diagnostic framework may generalize to more severe cases.

Recent studies have advanced dysarthria detection using machine- and deep-learning methods, but many rely on acoustic features that are hard to interpret clinically (for example, spectrogram- or Mel-frequency Cepstral Coefficients-derived inputs) and generate black-box models for clinical application (15, 16). Although some studies aim to improve interpretability, the resulting features remain abstract acoustic or phoneme-level representations, rather than clinically interpretable phonetic features (17). Speech recordings in many studies come from general speech repositories (e.g. TORGO) rather than sentences deliberately designed to accentuate dysarthric deficits (15). In contrast, our framework operates under a distinct diagnostic paradigm that emphasizes clinically and linguistically informed sentence design specific to dysarthria type and extracts simple meaningful syllable-error features via ASR. This keeps the inputs clinically readable and applicable for end users such as neurologists.

Transforming recorded speech into numerical syllable-error features obviates the need for complex preprocessing of raw acoustic data, for example speech noise filtering and signal segmentation. This simplification allows the use of a logistic regression classifier to predict spastic dysarthria without requiring an extensive sample size to achieve statistical power. Logistic regression is straightforward and transparent, providing clinical insight into the effect of consonant articulation on dysarthria detection. This highlights the value of clinically informed feature selection in simplifying data representation and supporting effective predictive modeling.

A key methodological strength of this study that differs from previous studies is the use of neuroimaging-confirmed corticobulbar lesions as the reference diagnostic standard, which we considered to be the ground truth, rather than relying solely on expert clinical diagnosis. As a result, a positive finding from our tool inherently reflects the presence of a corticobulbar lesion rather than just its clinical manifestation as dysarthria. This enhances its diagnostic utility by extending beyond symptomatic assessment to lesion localization, a critical aspect of neurological evaluation. While this implication could reduce reliance on advanced neuroimaging and enable more timely treatment, further work examining corticobulbar lesion laterality, number, and level is necessary to refine lesion-based diagnosis which lies beyond the scope of this study.

Another notable strength of this study is the use of Automatic Speech Recognition (ASR)-based analysis instead of subjective neurologist perception, which is prone to inconsistency, with poor intra-rater reliability (Cohen’s 𝛋 = 0.2–0.3) (6). ASR ensures objective and reproducible feature extraction while minimizing human bias. We selected Apple Siri™ ASR for its accessibility and reported Word Error Rate (WER) of under 10% in normal speech (18) but up to 70% in dysarthric speech (19, 20). Our assessment confirmed its ability to distinguish dysarthria from normal speech, with significant transcription error differences across all sentence types (see Fig. 2). ASR also demonstrated superior sensitivity in detecting mild dysarthria, offering finer severity grading compared to traditional methods, which clustered near the upper limit (Table 1).

Several limitations should be acknowledged. First, the case-control design limits generalizability of the findings, as the equal distribution of dysarthria and control participants does not reflect the true prevalence in clinical settings. This intentional oversampling of dysarthria cases may introduce bias in the estimation of sensitivity and specificity, necessitating cautious interpretation of these metrics.

Additionally, the case-control approach makes the study susceptible to selection bias, as the participant pool may not fully represent the entire spectrum of spastic dysarthria severity. These limitations underscore the need for external validation through prospective, population-based studies with consecutive recruitment of participants with suspected spastic dysarthria to establish real-world utility.

Second, two caveats from the ASR transcription process are noteworthy. One is that we observed lower Word Error Rate (WER) for common/high-frequency words than for uncommon words, which reduced the syllable-error gap between dysarthric and normal speech. Sentence design should therefore limit high-frequency words to preserve discriminability. Another caveat is that transcription performance varies by ASR platform. Platform-specific processing can shift WER and, in turn, downstream diagnostic metrics. Future studies should benchmark this paradigm across other platforms (e.g., Amazon Alexa™) using a similar methodological approach. In parallel, as general-purpose ASR improves at transcribing dysarthric speech, the resulting decrease in WER may diminish separability between dysarthric and normal speech, which warrants development of a dysarthria-aware ASR system optimized for dysarthria quantification.

Third, this proof-of-concept study focused exclusively on spastic dysarthria, given its high prevalence in clinical practice (2). However, diagnosing other types of dysarthria with different causative lesions remains challenging and requires methodological refinement. A practical next step to generalize the framework is to apply the same consonant-focused sentence set (LBP/LAP/VP/MP) to other types and examine whether the cross-sentence AUC pattern forms a reproducible diagnostic ‘signature’, since type-specific pathophysiologic and phonetic profiles likely shape distinct patterns of consonant articulation. For example, in flaccid/bulbar dysarthria caused by lower motor neuron lesions, increased hypernasality may introduce excessive nasal resonance to non-nasal consonants (all consonants except /m/, /n/, /ŋ/), potentially shifting the cross-sentence AUC profile toward sentences with a higher proportion of non-nasal consonants (e.g. the VP sentence). Alternatively, sentence design and error-scoring criteria could be modified to match type-specific phonetic deviations. For instance, incorporating rapid alternating syllable repetition (e.g., /pa-ta-ka/) into sentences for ataxic dysarthria due to cerebellar dysfunction could provide pathophysiology-aligned features alongside ASR-derived syllable-error measures. Building on these ideas, future research should develop classification approaches capable of differentiating dysarthria types and refining lesion localization—advancing beyond the current approach of distinguishing dysarthric from non-dysarthric speech.

Beyond diagnostic utility, the same approach may be well suited to longitudinal monitoring after corticobulbar lesion onset and during rehabilitation. Serial assessments using this paradigm could quantify within-patient improvement or relapse and characterize recovery trajectories to inform prognosis. Future studies should evaluate test-retest reliability, determine the clinically meaningful change in syllable-error scores, and compare their longitudinal trajectories with standard clinical scales and imaging follow-up to assess validity.

In conclusion, this study presents a novel approach to dysarthria lesion diagnosis, integrating phonetic-and-physiologic-based feature design, ASR-derived quantification of clinically meaningful transcription errors, and logistic regression modeling. This framework demonstrated the potential to diagnose spastic dysarthria with improved accuracy compared to expert perceptual assessment. The transformation of dysarthric speech into structured quantitative features not only enhances diagnostic performance but also translates computational results into clinical insight, revealing how phonetic factors, particularly consonant articulation, contribute to dysarthria identification.

Rather than relying on additional model complexities that require intensive computational power, our methodology emphasizes the significance of feature design, underscoring the integrative role of human expertise and artificial intelligence computation, an approach often described as ‘human craft, AI draft’. This paradigm fosters mutual learning, where clinical knowledge refines AI-driven analysis, while AI compensates for human diagnostic limitations, advancing dysarthria assessment in the digital era.

## Acknowledgements

This study would not have been possible without the contributions of the team members at the Chulalongkorn Cognitive Clinical & Computational Neuroscience (CCCN) Lab, whose invaluable advice supported the progress of this work. We would also like to express our deepest gratitude to Mrs. Chanunya Pollap, an expert speech-language pathologist at Chulalongkorn Hospital, for her significant contributions. Her expertise in linguistics and phonetics facilitated the critical design of phonetically-targeted sentences and their international phonetic descriptions to the speech assessment of each participant with dysarthria. We also sincerely appreciate the Chulalongkorn Stroke Center of Excellence for their support in case diagnosis and for facilitating speech data collection from the stroke ward. Finally, we are grateful to all patients and healthy participants at Chulalongkorn Hospital, Bangkok, Thailand, for their generous participation, which made this research possible and contributes to the advancement of digital medicine.

This research was supported by the Ratchadapisek Research Grant (Grant No. GA66/063). The funding primarily covered compensation for research participants and played no role in study design, data collection, analysis and interpretation of data, or the writing of this manuscript.

## Data availability Statement

Original clinical data, including de-identified speech audio files, demographic data, clinical information, and analyzed datasets (e.g., syllable error scores for all participants), are available upon request from the corresponding author and/or the Chulalongkorn Cognitive Clinical & Computational Neuroscience (CCCN) Center, Bangkok, Thailand due to the patient privacy reason. Data sharing is intended for research and academic purposes and is subject to patient privacy consideration under Thai Personal Data Protection Act (PDPA) and institutional privacy policies. Analysis code is publicly available at the following GitHub repository: https://github.com/CMKMITL/SpasticDysarthria-SyllableError-Analysis.git

## Author contributions

C.M. contributed to the initial drafting and final revision of the manuscript, the study concept and design, a major role in data acquisition and refinement, interpretation of the data, and preparation of figures and supporting information. C.C. contributed to the revision of the manuscript for intellectual content, initiated the study concept and design, and participated in data analysis and interpretation. K.K. contributed to data analysis and interpretation, and revised the manuscript. S.C. contributed to study design, data analysis, and manuscript revision. All authors reviewed the manuscript.

## Competing interests

The authors declare no competing interests.

*Large language models were used only to assist with language editing. The authors take full responsibility for the study design and conduct, data analysis, interpretation, and the manuscript content*.

## Supporting information captions

**S1 Fig | Intelligibility Score in Thai Dysarthria.** The figure presents the standard intelligibility assessment for Thai dysarthric speech. A speech-language pathologist conducted a perceptual evaluation in which dysarthric patients were instructed to pronounce 36 Thai words clearly. The assessor assigned a score based on the number of intelligible words, with a maximum possible score of 36.

**S2 Fig | Permutation-based null distributions of AUC scores for each sentence model.**

Histograms show the null distribution of test AUC values generated from 10,000 label permutations for logistic regression models based on individual sentences (LBP, LAP, VP, and MP) and ‘combined’ sentence. The red dashed vertical line indicates the median AUC value from the original, non-permuted analysis. *P*-values denote the proportion of permuted AUCs equal to or greater than median AUC (one-sided).

**S1 Table | Automatic Speech Recognition transcription results.** Table displays a sample of ASR-transcribed sentences, segmented into syllables for 10 selected participants (out of 74 participants). Incorrectly transcribed syllables (highlighted in pink) were simply counted to determine error scores based on predefined criteria.

**S2 Table | Shapiro-Wilk Normality test of syllable error scores.**

Table presents normality test results for error scores across all sentences. Most distributions deviated significantly from normality (*p* < 0.05), except for the VP sentence in the spastic dysarthria group.

**S3 Table | Mann-Whitney U Test for syllable error scores by sentence.** Table presents the median and interquartile range (IQR) of error scores, along with mean rank, sum of ranks, test statistic (U), and *p*-values for each sentence, comparing the spastic dysarthria and control groups.

